# Entity-Aware Generation of Synthetic Clinical Progress Notes for Prostate Cancer using Large Language Models

**DOI:** 10.64898/2026.06.12.26355166

**Authors:** Álvaro Rey-Blanes, Javier Veredas-Morente, Francisco J. Moreno-Barea, Francisco J. Veredas

## Abstract

**Objectives:** This study investigates large language models (LLMs) for clinical entity projection across substantial textual transformation. Specifically, we evaluate whether entities annotated in Spanish prostate cancer case reports can be preserved and explicitly projected when the source narratives are transformed into hospital-style clinical progress notes. Entity projection is treated as a generation-driven task, allowing paraphrase, condensation and narrative reorganisation, providing that clinically relevant entities remain recoverable as structured annotations.

**Methods:** A corpus of 109 Spanish prostate cancer case reports was annotated using a silver-standard pipeline combining Spanish biomedical named-entity recognition with rule-based prostate-specific antigen (PSA) and Gleason extractors. The resulting silver-standard annotations were validated on a subset of generated notes against a gold-standard consensus produced by medical experts in prostate cancer. Four LLMs were evaluated for note generation and entity projection: GPT-5.4 Nano, Qwen 3.5:35B-A3B, GLM5 and Claude Sonnet 4.6. Entity-to-Entity (E2E) generation used XML-annotated cases as RAG-supported input, whereas Text-to-Entity (T2E) generation required models to generate and annotate notes directly from plain text cases. Zero-shot and few-shot prompting were tested. Projection quality was measured using precision, recall and F1-score, and complemented by LLM-as-a-judge evaluation using Kimi K2.6.

**Results:** E2E consistently outperformed T2E, indicating that explicit entity-enriched input substantially facilitates entity preservation and localisation. GLM5 achieved the best E2E zero-shot result (F1 = 0.915), followed by Claude Sonnet 4.6 (F1 = 0.896). In T2E, few-shot prompting improved performance, with Claude Sonnet 4.6 reaching the highest score (F1 = 0.718). *Age*, *Gleason*, *Disease*, *Procedure*, *Duration* and negation-related entities were robustly projected, whereas *PSA* and *Dose* showed less stable behaviour.

**Conclusion:** LLMs can generate clinically plausible synthetic prostate cancer evolution notes while preserving a substantial proportion of source entities, particularly when explicit semantic annotations are provided as input. However, the lower and more variable performance observed in T2E highlights the difficulty of jointly generating clinical narratives and projecting entities without source-side information, especially for numerical and measure-related entities.

## 1 Introduction

### 1.1 Medical Progress Notes and Clinical Case Reports

Clinical documentation is a central component of modern healthcare, particularly in specialities where patient management depends on the interpretation of longitudinal information. In electronic health records (EHRs), clinical progress notes, also known as evolution notes, provide a narrative account of the patient’s status, relevant clinical events, diagnostic findings, therapeutic decisions and subsequent changes over time. Their function is especially important in continuity of care [12], where clinicians must reconstruct the patient trajectory from information produced across multiple encounters. This need is particularly evident in complex oncological conditions such as prostate cancer, in which urologists and oncologists routinely integrate pathological findings, imaging results, laboratory values, treatment history and disease progression to support clinical decision-making [30].

Clinical notes are essential for both care continuity and information management. By bringing together temporally dispersed and heterogeneous information, including diagnostic procedures, therapeutic interventions, and relevant clinical events, they provide a shared narrative that supports communication and handover between healthcare professionals. This function is closely associated with safer and higher-quality care, as it reduces the likelihood of information loss, ambiguity or misinterpretation [50]. At the same time, the production of clinical documentation represents a substantial workload for clinicians, who often spend a considerable proportion of their working time on record-keeping rather than direct patient care [46, 45]. This documentation burden has been recognised as a contributor to healthcare inefficiency, clinician burnout and reduced productivity [46]. These challenges are further intensified by the considerable variability with which medical evolution notes are written across clinicians, including differences in terminology, structure, level of detail and writing style. Such heterogeneity limits interpretability and complicates downstream computational processing. Moreover, because the manual generation of evolution notes is time-consuming and difficult to scale in high-demand clinical environments, maintaining consistent and high-quality documentation becomes increasingly challenging as patient volume increases [45].

Biomedical case reports also describe individual patient trajectories and constitute an important form of published clinical knowledge [16]. However, their communicative purpose differs from that of routine clinical documentation. Case reports are usually written retrospectively and are often selected because the case is rare, educationally valuable or clinically relevant. Consequently, they tend to present the clinical course in a more complete, coherent and explanatory manner, including diagnostic reasoning, treatment decisions and outcomes.

These differences have direct implications for the structure and usability of both text types. Progress notes are commonly produced under time pressure and within the constraints of routine care, which often leads to fragmented narratives, abbreviations, variable levels of detail and heterogeneous organisational patterns [25]. Case reports, by contrast, are generally more polished and temporally ordered, since they are written after the clinical trajectory has unfolded. As a result, EHR notes better reflect the complexity and variability of everyday clinical practice, whereas case reports provide cleaner and more interpretable narratives, albeit shaped by retrospective selection and publication bias.

Both clinical progress notes and biomedical case reports contain a high density of clinically relevant entities, including diseases, diagnoses, signs and symptoms, findings, procedures, drugs, laboratory values, anatomical references and temporal expressions. The automatic identification of these mentions is commonly formulated as a named entity recognition (NER) task and constitutes a fundamental step for many downstream clinical NLP applications, such as information extraction, cohort identification, clinical decision support and semantic indexing of patient trajectories. However, NER in specialised biomedical domains remains challenging because entity mentions are highly domain-dependent, context-sensitive and often expressed through specialised terminology, lexical variation, nested concepts or ambiguous abbreviations [26]. These difficulties are amplified in non-English clinical domains [17], such as Spanish prostate cancer narratives, where fewer annotated resources, terminological variants and institution-specific writing conventions limit the portability of general-purpose or out-of-domain models. Although case reports usually provide more explicit and coherent descriptions of the clinical course, routine progress notes pose an even more complex scenario for entity recognition due to their fragmented structure, telegraphic style, frequent use of abbreviations, implicit references, incomplete sentences and substantial variability across clinicians.

In this work, we address this problem from the perspective of synthetic clinical text generation and entity projection. Large language models (LLMs) offer a promising way to generate synthetic clinical progress notes from biomedical case reports, thereby helping to alleviate the scarcity of accessible, privacy-preserving and clinically realistic EHR data needed to train and evaluate AI systems intended for real-world clinical documentation [19, 54]. However, generating realistic clinical prose is not sufficient in itself: the resulting notes must also preserve the clinically salient information contained in the source case [52]. We therefore investigate whether LLMs are able not only to transform a case report into a synthetic progress note, but also to faithfully project the entities identified in the original text into the generated note. This setting allows us to evaluate entity preservation, omission, transformation and hallucination during generation, while also producing entity-enriched synthetic notes that can mitigate the difficulty of performing NER directly on heterogeneous clinical documentation. In particular, we analyse entity-aware generation strategies in which retrieval-augmented generation (RAG) [3] is used to enrich the source cases with semantic information about their clinical entities, with the aim of assessing whether this additional supervision improves the projection of relevant entities into the generated prostate cancer progress notes.

### 1.2 Large Language Models in Healthcare

LLMs have become increasingly influential in biomedical and clinical natural language processing (NLP). Built mainly on transformer-based architectures and trained on large-scale textual corpora, these models are able to process, interpret and generate complex natural language with a level of fluency and contextual awareness that makes them particularly suitable for clinical free-text applications [47]. Their relevance is especially clear in healthcare settings, where a substantial proportion of patient information remains embedded in unstructured narratives such as EHRs, discharge summaries, radiology reports, clinical case reports and progress notes.

One of the most prominent uses of LLMs in this domain is clinical summarisation. By producing abstractive summaries from long, heterogeneous and context-rich documents, these models can help condense clinically relevant information into more concise and coherent narratives [5]. This capability is particularly valuable when longitudinal patient information must be synthesised into structured evolution notes or other forms of clinical documentation [27]. Beyond summarisation, LLMs have also been explored for clinical question answering, where they are used to retrieve, interpret, and synthesise information from patient records or biomedical literature in response to clinically oriented queries [62]. In this context, they have been investigated as tools for supporting information retrieval, clinical reasoning and decision-making processes [38].

Recent work has also examined the use of LLMs to reduce the burden associated with clinical documentation. These systems can generate high-fidelity clinical narratives, including progress notes, discharge summaries and case descriptions, from heterogeneous inputs such as clinician-patient conversations, structured records or previously documented clinical events [7]. This line of research directly addresses the need to streamline documentation workflows by automating parts of the clinical writing process while preserving medically meaningful content [24].

Despite their potential, the use of LLMs in clinical environments raises important safety and reliability concerns. A major limitation is the risk of hallucination, whereby the model generates information that is not supported by the input data. In healthcare, such errors may result in fabricated diagnoses, inaccurate laboratory values, incorrect treatments or unsupported clinical events, all of which may compromise the reliability of the generated text. In addition, LLM outputs may contain internal contradictions, temporal inconsistencies or violations of clinical logic. These limitations are especially relevant in longitudinal documentation, where the correct ordering of events and the preservation of clinical coherence are essential for interpretability and safe downstream use [4].

### 1.3 Current Problem and Contribution

The main objective of this work is to evaluate the ability of LLMs to generate synthetic clinical evolution notes from prostate cancer case reports while preserving and explicitly projecting the clinically relevant entities contained in the source narratives. Previous work has shown that LLMs can transform published case reports into synthetic evolution notes that resemble real-world EHR documentation and retain a substantial proportion of the clinical information present in the original cases [54]. However, generating narratives that are plausible in form and style is only one part of the problem. For synthetic clinical text to be useful in downstream clinical NLP applications, such as NER, information extraction or corpus construction, the generated notes must also preserve the core clinical content of the source case, and relevant entities must be explicitly represented and correctly localised in the generated narrative. In this study, we therefore analyse entity projection as a way to assess the extent to which LLMs maintain essential clinical information when converting prostate cancer case reports into synthetic evolution notes following narrative conventions observed in routine hospital EHR documentation.^1^ This is a non-trivial task, since LLM-generated notes may preserve entities literally, reformulate them using equivalent expressions, normalise them, reorganise them temporally, omit them or introduce unsupported information. In prostate cancer, this challenge is particularly relevant because key entities include diagnoses, procedures, treatments, disease status, PSA values, Gleason scores, staging information and clinically significant temporal events.

Moreover, the main added value of this work lies in treating synthetic clinical notes as structured and auditable NLP resources rather than as plain generated text. In this context, entity projection provides a mechanism for preserving relevant information between the original clinical case and the generated evolution note. Instead of simply applying an entity recogniser to the synthetic text, the proposed approach evaluates whether source entities can be automatically located in the generated narrative and whether their clinical meaning is preserved after transformation. This distinction is important because a synthetic note may be fluent and clinically plausible while still omitting, distorting, or relocating relevant information from the original case.

Building on this objective, the generated and projected notes must satisfy three essential requirements. First, the generated narrative must remain faithful to the source case report, avoiding unsupported additions, omissions of critical information and clinically relevant distortions. Second, the synthetic note must preserve the temporal and clinical logic of the patient trajectory, ensuring that diagnostic findings, treatments, disease progression and outcomes are presented in a coherent sequence. Third, the projected entities must be accurate and clinically meaningful, with correct span localisation and entity categorisation whenever the corresponding information is present in the generated text.

These requirements make the task more demanding than conventional clinical summarisation or standard NER. The model must not only generate fluent and realistic clinical language, but also preserve the clinical content required for entity-level traceability. At the same time, the projection process must account for the variability introduced by abstractive generation, including paraphrases, lexical variation, reordered information and partial mention reformulation. Therefore, this work addresses a methodological gap between synthetic clinical text generation and the construction of annotated resources for clinical NLP.

The present work makes the following main contributions:

- First, we propose a framework for generating synthetic Spanish prostate cancer evolution notes and automatically projecting clinically relevant entities from the source case reports onto the generated narratives. This allows the resulting notes to be treated not only as synthetic text, but also as entity-enriched clinical NLP resources.
- Second, we compare two entity-aware generation settings to analyse the effect of retrieval-based contextual support on clinical fidelity, entity preservation and projection quality: Entity-to-Entity (E2E) Generation uses RAG-supported, entity-enriched clinical cases as input to generate entity-enriched clinical progress notes, whereas Text-to-Entity (T2E) Generation generates entity-enriched clinical progress notes directly from plain-text clinical cases, without retrieval-based entity enrichment.
- Third, we analyse the preservation and recoverability of key medical entities, including diagnostic findings, procedures, treatments, PSA values, Gleason scores, disease status, staging information and temporal clinical events while we evaluate the projected annotations in terms of entity localisation, clinical consistency and projection errors, distinguishing between exact preservation, reformulation, omission, boundary mismatch and unsupported generated content.
- Fourth, we provide a methodological contribution for the construction of synthetic annotated clinical corpora, showing how LLM-generated notes can be enriched with projected entities while maintaining traceability to the original clinical case.

## 2 Related Work

### 2.1 Clinical Text Summarisation

summarisation has become an important line of work in clinical NLP because patient information is rarely concentrated in a single document. Instead, it is dispersed across progress notes, diagnostic tests, imaging reports, procedures, treatments and follow-up records, increasing the effort required to reconstruct the relevant clinical trajectory. This contributes to the broader documentation burden in healthcare, where a substantial proportion of physicians’ working time (up to 49.2%) is devoted to EHR-related and desk work [58]. Earlier summarisation systems mainly followed extractive strategies, selecting relevant passages from the source documents and preserving a strong link with the original text [43]. Although this improves factual grounding, it provides limited support for tasks that require temporal integration, clinical abstraction and synthesis of information distributed across multiple encounters [**?**, 11, 51].

For this reason, recent approaches have increasingly explored abstractive or hybrid methods capable of reorganising and reformulating clinical content into coherent summaries [55]. Such methods are better suited to longitudinal records and patient trajectory summarisation [27], but they also introduce risks related to factual accuracy, temporal consistency and preservation of clinically relevant details, particularly when the input text is noisy, incomplete or stylistically heterogeneous [64]. These issues are directly relevant to this study, where summarisation is not the final objective but an intermediate step: prostate cancer case reports are transformed into synthetic evolution notes that must preserve enough clinical information to enable reliable downstream entity projection.

LLMs have shifted clinical summarisation towards generative methods, mainly through prompting and fine-tuning. Prompting uses task-specific instructions or examples to guide pre-trained models without additional training, making it flexible and less dependent on annotated data. Fine-tuning, in contrast, adapts the model to clinical or domain-specific corpora, which may improve performance but requires more data, computation and control of privacy-related risks [20]. Reasoning-based prompting strategies, such as chain-of-thought, have also been explored to improve coherence and structure in generated text [63]. However, in clinical summarisation these techniques require caution, since additional reasoning steps may introduce unsupported inferences or factual inconsistencies when strict grounding in the source document is required.

### 2.2 Generation of Synthetic Clinical Text

The development of clinical NLP resources is strongly constrained by the limited accessibility of real patient records. Privacy regulations such as HIPAA and GDPR restrict the sharing and reuse of clinical data, and even de-identified notes may still be exposed to privacy risks, including membership inference attacks (MIA)[14, 23, 56]. In this context, synthetic clinical text generation has become an increasingly relevant strategy for producing shareable corpora while reducing direct dependence on sensitive hospital data [22, 37, 56]. Early approaches explored the generation of short clinical narratives or record fragments using encoder-decoder models, GAN-based architectures, LSTMs with differential privacy and transformer-based systems applied to domains such as chief complaints, EHRs, MIMIC-III notes and mental health documentation [31, 21, 40, 23].

The field has since shifted towards large pre-trained language models, which have shown greater capacity to generate fluent and clinically plausible narratives. Comparative studies have reported the advantage of models such as GPT-2 over earlier neural architectures for generating clinical sections useful in downstream NER tasks [32]. More recent work has explored high-fidelity synthesis with LLMs, including literature-derived synthetic notes, guideline-conditioned discharge summaries, task-specific data augmentation for medical coding and specialised applications in radiology using both proprietary and open-source models[28, 13, 15, 6, 37, 48].

Recent work has increasingly moved towards guided synthetic generation, where LLM outputs are conditioned by clinical guidelines, predefined structures or specific downstream objectives. Ellershaw et al. used Royal College of Physicians guidelines to generate structured discharge summaries [13], while other studies have explored synthetic clinical notes as task-specific augmentation data for automated medical coding and ICD assignment [15, 6]. In radiology, local open-source LLMs have also been evaluated for clinically constrained tasks, including fracture identification and the conversion of dictated reports into structured templates [37, 48]. This line of work shows that synthetic clinical text generation is increasingly being framed around controlled outputs with explicit clinical or computational utility.

In the Spanish clinical NLP setting, synthetic text generation is gaining relevance as a strategy to reduce the scarcity of non-English clinical resources. Platas et al. proposed a hybrid approach in which structured real-world data from liver cancer cases were used to generate and automatically annotate Spanish Computed Tomography (CT) reports, showing that the inclusion of synthetic examples improved NER performance in small-data scenarios [49]. Similarly, Vakili et al. investigated data-constrained synthesis for de-identification in Swedish and Spanish, demonstrating that moderately sized LLMs can be adapted with limited in-domain data to generate useful synthetic corpora for NER tasks [61]. These studies support the use of synthetic generation as a practical route for expanding clinical NLP datasets in Spanish and other low-resource languages. In our previous work, we evaluated the generation of realistic Spanish prostate cancer progress notes from published case reports using LLMs, showing that the resulting synthetic notes can resemble hospital-based EHR documentation while preserving relevant clinical information from the source narratives [54]. The present study builds on that work by moving from narrative realism and general clinical consistency towards explicit entity projection, assessing whether clinically relevant entities in the source cases are preserved, transformed and correctly localised in entity-enriched synthetic progress notes.

### 2.3 Clinical Entities Projection

Entity-level preservation has become an increasingly relevant issue in clinical text generation, since a generated note may be fluent and clinically plausible while still omitting, altering or misplacing important medical concepts. In clinical summarisation, recent work has shown that explicitly modelling salient entities can improve both coverage and faithfulness. Adams et al. proposed SPEER, a sentence-level planning strategy in which relevant entity spans are marked in the source notes and retrieved during summary generation, allowing the model to track which clinical concepts are being used in each generated sentence [1]. This line of work is closely related to entity projection, as it treats entities not only as output labels but also as anchors for controlling and auditing the generation process.

Other studies have explored LLMs as tools for clinical information extraction from specialised medical documents. Choi et al. developed prompts to extract relevant clinical factors from pathology and ultrasound reports in breast cancer [10], while CORAL introduced an expert-curated oncology resource designed to evaluate language models on detailed oncological information, including entities, modifiers and relationships [60]. More broadly, recent work has investigated LLM-based NER and entity linking in multilingual biomedical settings [39], as well as the extraction of named entities from summarised medical text [57]. These contributions show that clinical entities are increasingly being used to structure, evaluate and control LLM-based clinical NLP systems.

However, most existing approaches focus either on extracting entities from original clinical documents, using entities to guide summarisation, or evaluating LLMs on information extraction tasks. The specific problem addressed in this work is different: clinical entities are projected from source case reports onto synthetic evolution notes generated by LLMs. This requires identifying whether source entities are preserved in the generated text, whether they appear literally or through reformulation, and whether their clinical meaning and span boundaries remain valid after generation. Therefore, the task combines synthetic clinical text generation, entity-level traceability and automatic annotation transfer.

To the best of our knowledge, automatic projection of clinically relevant entities onto LLM-generated synthetic Spanish oncology evolution notes remains underexplored. This study addresses that gap by analysing entity projection under two generation settings—with RAG (E2E) and without RAG (T2E)—, allowing us to evaluate whether retrieval support improves not only narrative generation but also entity preservation, localisation and downstream annotation quality.

## 3 Material and Methodology

### 3.1 Data and Silver-Standard Entity Recognition Pipeline

The source material for this study corresponds to a curated dataset of 109 Spanish-language prostate cancer case reports made publicly available in Zenodo by Rey-Blanes et al. [53]. The dataset contains individual patient cases extracted from the biomedical literature and provides detailed clinical narratives covering presentation, diagnostic work-up, therapeutic management, disease evolution and outcomes. Its composition makes it suitable for controlled synthetic clinical text generation, as the documents offer sufficiently rich and structured descriptions of patient trajectories while remaining publicly accessible. Since the corpus is derived from published case reports rather than private hospital EHRs, it also enables the evaluation of both local and cloud-based LLMs under reduced privacy constraints.

To obtain the entity-level reference used throughout this study, we implemented a hybrid silver-standard entity recognition pipeline. This pipeline was applied in a consistent manner to both the source prostate cancer case reports and the synthetic evolution notes generated under the E2E and T2E settings. Its purpose was twofold. First, it provided an automatically derived inventory of clinically relevant entities in the source narratives, which served as the basis for the projection process. Second, it generated the silver-standard annotations used to compute entity-level metrics on the generated notes, allowing the comparison of entity coverage, preservation and projection quality across generation strategies.

The annotation pipeline combined neural NER models with rule-based extractors. This design was adopted because the study required broad clinical coverage, including diseases, symptoms, procedures, drugs, temporal expressions and negation/speculation phenomena, while also capturing prostate-cancer-specific variables such as PSA values and Gleason scores. The same pipeline was therefore used as a common reference mechanism for analysing both the original clinical information and its representation in the synthetic texts. In this setting, the silver standard should not be interpreted as manually validated gold-standard annotation, but as a systematic and reproducible automatic reference layer for downstream comparison.

The first component of the pipeline consisted of six Spanish biomedical-clinical NER models developed by the *Barcelona Supercomputing Center (BSC)* and based on the *BSC-NLP4BIA/PlanTL-GOB-ES* model family. These models derive from `bsc-bio-ehr-es`, a RoBERTa-based Spanish biomedical-clinical encoder pretrained on large-scale biomedical text and EHR data [9]. The selected downstream token-classification models were used to cover complementary clinical entity categories:

- `BSC-NLP4BIA/bsc-bio-ehr-es-livingner-species`: model fine-tuned within the LivingNER framework to identify species and organism mentions in biomedical and clinical texts [41].
- `BSC-NLP4BIA/bsc-bio-ehr-es-livingner-humano`: model fine-tuned for the detection of human mentions, also derived from LivingNER resources [41].
- `BSC-NLP4BIA/bsc-bio-ehr-es-medprocner`: model fine-tuned for the recognition of medical procedures in Spanish clinical narratives using MedProcNER resources [34].
- `BSC-NLP4BIA/bsc-bio-ehr-es-symptemist`: model fine-tuned to identify symptoms, signs and clinical findings using SympTEMIST resources [35].
- `BSC-NLP4BIA/bsc-bio-ehr-es-distemist`: model fine-tuned for disease mention detection using DisTEMIST resources [42].
- `PlanTL-GOB-ES/bsc-bio-ehr-es-pharmaconer`: model fine-tuned for pharmacological substances, compounds and related biomedical mentions using PharmaCoNER resources [18].

In addition to the BSC models, three models from the MedSpaNER family were incorporated to capture semantic phenomena that are particularly relevant for interpreting clinical narratives [8]. These models expanded the silver-standard layer beyond conventional entity recognition by adding medication attributes, temporal information and negation/speculation annotations:

- `medspaner/roberta-es-clinical-trials-cases-medic-attr`: RoBERTa-based model used to identify medication-related attributes, including dosage, route, form and other drug-associated information.
- `medspaner/xlm-roberta-large-spanish-trials-cases-temp-ent`: XLM-RoBERTa-large-based model used to detect temporal entities, including dates, ages, durations, frequencies and other time expressions.
- `medspaner/roberta-es-clinical-trials-cases-neg-spec`: RoBERTa-based model used to recognise negation and speculation phenomena, including negation cues, negated concepts, speculation cues and speculated clinical events [36].

To complement the neural models, two rule-based extractors were implemented for prostate-cancer-specific variables that are essential for clinical interpretation but frequently appear as compact numerical patterns or abbreviated expressions. The PSA extractor identified prostate-specific antigen values followed by common unit variants such as `ng/mL`, allowing both decimal commas and decimal points. The Gleason extractor captured frequent scoring formats, including primary and secondary pattern combinations such as `3+4`, total-score mentions such as `Gleason 7`, and combined formulations such as `Gleason 7 (3+4)` or `Gleason 3+4=7`. These rules were incorporated to improve the recall of clinically critical variables for prostate cancer staging, prognosis, progression assessment and treatment interpretation.

The combined output of the neural and rule-based components constituted the silver-standard annotation framework used in the experiments. Applied to the source reports, it defined the reference set of clinical entities to be projected. Applied to the generated notes, it provided the entity annotations required to compute silver-standard metrics, including entity coverage, entity-type distribution, preservation of prostate-cancer-specific variables and differences between RAG-based and non-RAG generation. Examples and definitions for each label can be found in Appendix A.

### 3.2 Experiments and Prompting

E2E and T2E were designed as two related synthetic generation settings for prostate cancer progress notes, both aiming to transform an original case report into a concise hospital-style clinical narrative while preserving information relevant to urological management. In both settings, the generated text was required to retain clinically meaningful information on diagnosis, staging, treatment, PSA, Gleason score, procedures, temporal evolution and complications, while avoiding unsupported inference or the addition of new clinical facts. The expected style was deliberately closer to real-world Spanish hospital documentation than to the narrative style of published case reports: short, information-dense sentences, frequent clinical abbreviations, minimal explanatory language and a chronological structure centred on diagnosis, treatment, progression, complications and final status. The main difference between the two settings lies in the role of entity annotation during generation. In E2E, the model was provided with a fully XML-annotated clinical case as RAG and was asked to generate an equivalent clinical note while preserving a comparable XML-tagged structure. Thus, the model could rely on pre-existing entity annotations in the input. In T2E, the input case was not pre-annotated in the same way; instead, the model had to both generate the rewritten clinical note and identify the clinically relevant entities using the predefined XML-like tag set. T2E therefore combined note generation with entity recognition and annotation, making it a more demanding setting. This distinction is central to the interpretation of the results: E2E mainly evaluates whether models can preserve and reformulate already marked clinical information, whereas T2E evaluates whether they can infer, select and annotate the relevant clinical entities while generating the synthetic note. Complete prompts (original in Spanish and its English translation) can be observed in Appendix B.

### 3.3 Validation of the Silver-Standard for Subsequent Experiments

Before using the silver-standard annotations as the reference layer for the subsequent projection experiments, we assessed their agreement with expert annotations on a subset of 20 generated progress notes. This subset was obtained from the GLM5-E2E-few-shot setting, described in Section 3.4. These notes were independently reviewed by two medical experts, and a consensus version was produced and used as the reference annotation for this validation step. The purpose of this analysis was not to evaluate the performance of a specific evaluation setting, but to determine whether the silver standard was sufficiently aligned with expert consensus to serve as a scalable reference label for the remaining experiments.

As shown in Table 1, the silver standard showed a high degree of agreement with the expert consensus, reaching an F1-score of 0.81. Its agreement to the consensus was nearly identical to that obtained by the predictions from the GLM5-E2E-few-shot setting, which achieved an F1-score of 0.811. The silver standard showed slightly higher precision, whereas the GLM5-E2E-few-shot predictions showed slightly higher recall.

**Table 1:** Agreement of the silver standard and the predictions from the GLM5-E2E-few-shot setting with a two-expert consensus on 20 generated progress notes. Entity matches required the same label and either the same start or end boundary.

| Comparison | Precision | Recall | F1-score | Accuracy |
| --- | --- | --- | --- | --- |
| Silver standard vs. expert consensus | 0.865 | 0.761 | 0.810 | 0.680 |
| GLM5-E2E-few-shot vs. expert consensus | 0.859 | 0.769 | 0.811 | 0.682 |

This close agreement suggests that the silver standard captures substantially the same general annotation space as the manually reviewed expert consensus. Therefore, it can be used as a suitable and scalable reference annotation layer for evaluating entity projection quality across the remaining experiments and models. In total, the expert consensus, the silver standard, and the GLM5-E2E-few-shot predictions contained 782, 688, and 700 entities respectively.

**Figure 1:**
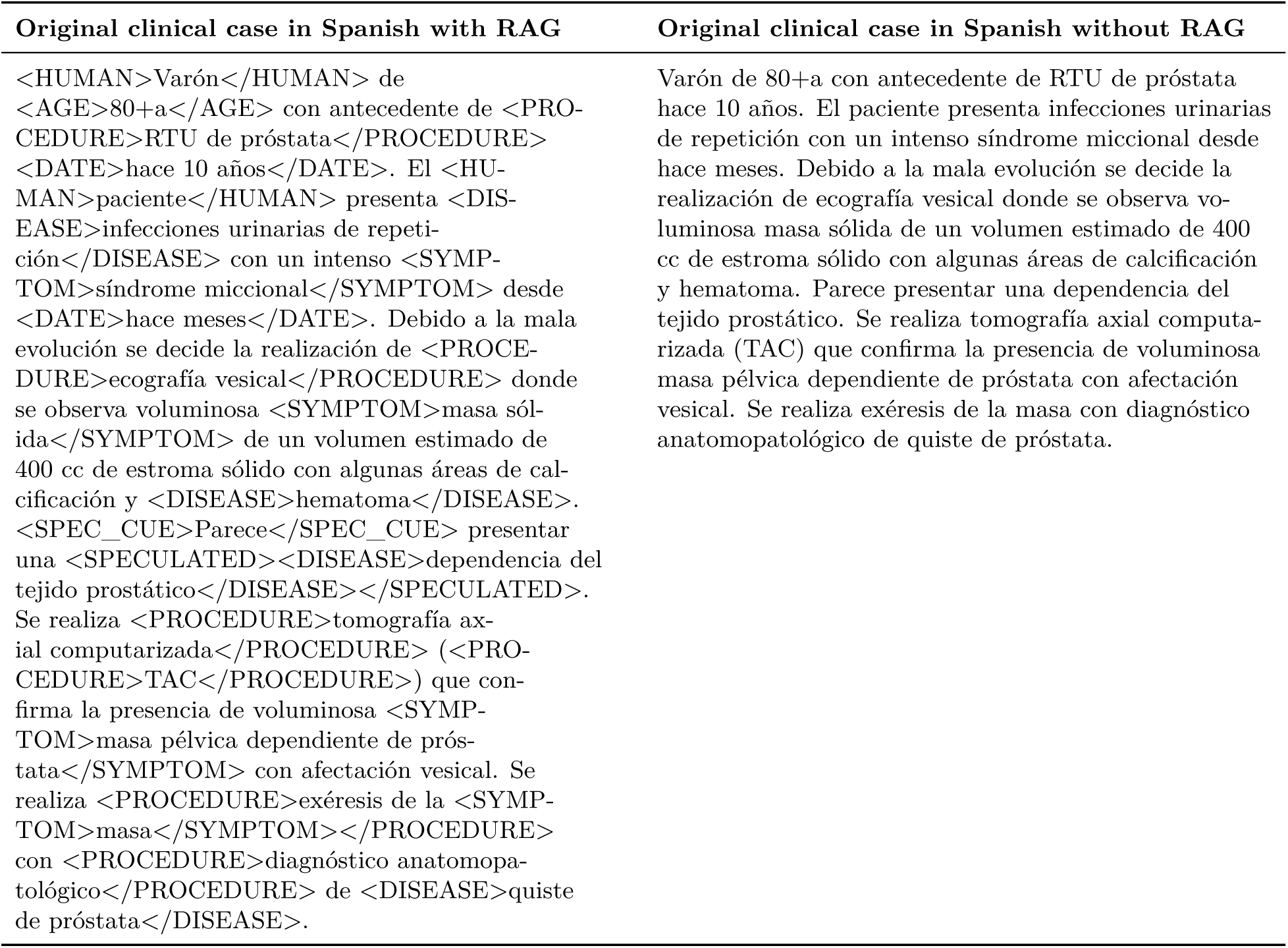
Qualitative example for patient 2 illustrating the main observable difference between E2E and T2E. The left column shows the original clinical case with XML-style entity annotations used by the RAG-based E2E configuration, whereas the right column shows the original clinical case for T2E with the same clinical content expressed as plain text. See complementary Figure C1 for English version

### 3.4 LLMs Used for Clinical Note Generation and Entity Projection

This study evaluates a heterogeneous set of LLMs for the generation of realistic synthetic clinical evolution notes and for the subsequent structured assessment of the generated outputs. The selected generation LLMs include GPT-5.4 Nano, Qwen 3.5:35B-A3B, GLM5 and Claude Sonnet 4.6, representing different deployment strategies, model families and access modalities. Each generation model was evaluated under the two entity-aware generation strategies proposed in this work: Entity-to-Entity (E2E) Generation and Text-to-Entity (T2E) Generation. For both strategies, we considered two prompting configurations: a zero-shot setting, in which the model received only the task instructions, and a few-shot setting, in which the prompt was enriched with two manually crafted examples prepared by a clinical expert to illustrate the expected input–output transformation and entity annotation format. This resulted in a factorial experimental design in which each generation model was tested across both generation strategies and both prompting settings.

In addition, Kimi K2.6 was used as an independent LLM-as-a-judge evaluator. This separation between generation and evaluation was adopted to reduce model-specific bias and to provide a more robust assessment of the generated clinical narratives. A summary of the models included in the study is provided in Table 2.

- **GPT-5.4 Nano (OpenAI)** was included as a proprietary cloud-based model accessed through an API. Since it is a closed-source system, architectural details and parameter count are not publicly disclosed. It was selected because of its efficient inference profile, large-context capabilities and suitability for instruction-following generation tasks requiring controlled clinical reformulation.
- **Qwen 3.5:35B A3B** was used as an open-weights model for local inference. Its A3B configuration provides an optimised deployment setting for efficient generation while preserving strong multilingual and reasoning capabilities. Local execution offers greater control over inference parameters and reduces exposure of input data to external services. In this study, the model was deployed on two dedicated GPU instances with 32GB of VRAM, achieving a mean latency of ≈ 7 seconds per patient and a throughput of ≈ 75 tokens per second. Its instruction-following capacity makes it suitable for constrained clinical generation, where the output must preserve source information, relevant entities and temporal coherence [2].
- **GLM5** was selected as a large multilingual general-purpose model based on a Mixture-of-Experts architecture. This design enables dynamic routing through specialised expert components, improving scalability for complex generation tasks. Given its large parameter scale, inference was performed through cloud access. The model was included to assess the behaviour of a high-capacity multilingual system in the generation of clinically constrained prostate cancer evolution notes [65].
- **Claude Sonnet 4.6 (CS4.6 - Anthropic)** was included as a proprietary cloud-based generation model. It was selected to complement the comparison with another closed-source system with strong instruction-following and long-context capabilities. In this study, Claude Sonnet 4.6 was used to generate synthetic clinical evolution notes under the same task constraints as the other generative models, allowing comparison across proprietary, open-weights, local and cloud-based systems.
- **BioMistral 7B** was considered as a biomedical domain-adapted baseline [29]. Its inclusion was intended to examine whether biomedical pre-training provided advantages over larger general-purpose models in this constrained generation task. However, its outputs did not reach the minimum quality threshold established for the study, particularly in terms of clinical fidelity and structural adequacy. For this reason, BioMistral 7B was excluded from the main comparative analysis.
- **Kimi K2.6** was used as the independent LLM-as-a-judge evaluator. Its role was not to generate clinical notes, but to provide structured assessments of the generated outputs according to the evaluation criteria defined in the study. Using a separate evaluator model ensured that the quality assessment was not performed by any of the models involved in generation, thereby reducing self-evaluation bias [44].

**Table 2:** LLMs used for synthetic clinical progress note generation, entity projection and LLM-as-a-judge evaluation.

| Model | Role | Access | Notes | Parameters |
| --- | --- | --- | --- | --- |
| *Generation models* |  |  |  |  |
| Claude Sonnet 4.6 | Generator | Cloud | Proprietary general model | Undisclosed |
| GLM5 | Generator | Cloud | Large open source MoE model | 744B |
| GPT-5.4 Nano | Generator | Cloud | Proprietary general model | Undisclosed |
| Qwen 3.5:35b | Generator | Local | Open-weights, controlled inference | 35B, A3B active |
| *Evaluation model* |  |  |  |  |
| Kimi K2.6 | LLM-as-a-judge evaluator | Cloud | Independent evaluator model | 1.04T |

*Note.* BioMistral 7B was additionally considered as a biomedical domain-adapted baseline. However, its generated outputs did not satisfy the minimum quality criteria established for the task and were consequently excluded from the main comparative analysis.

### 3.5 Evaluation Framework

The clinical adequacy of the generated notes was assessed according to the evaluation framework previously defined for generated progress notes [54]. This framework combines a preliminary safety screening with a structured quality assessment based on criteria adapted from the *PDQI-9* instrument [59] and 5-point Likert-scale scoring [33]. The safety screening identifies outputs with critical clinical failures, including unsupported clinical assertions, erroneous diagnostic or therapeutic information, or contradictions that compromise the interpretation of the disease course. The quality criteria considered both fidelity to the source case, including temporal correctness, factual precision and exhaustiveness, and intrinsic document quality, including clinical usefulness, organisation, comprehensibility, conciseness, synthesis and internal consistency. In the present study, this clinical evaluation provides methodological context for the generated notes, whereas the primary quantitative analysis focuses on entity-level agreement between projected annotations and silver-standard annotations in the synthetic narratives.

Entity-level evaluation was performed by comparing BRAT .ann files from the reference annotations and the evaluated annotations, paired by document identifier. Only text-bound annotations were considered, while discontinuous spans were excluded from the analysis. This restricted the evaluation to explicit entity mentions with standard span offsets.

A predicted entity was considered a true positive (TP) when it had the same entity label as a reference annotation and shared either the same start offset or the same end offset. This relaxed boundary criterion was used to tolerate minor span-boundary variation while still requiring agreement in entity type and positional alignment. Matching was one-to-one, so each reference annotation and each predicted annotation could be matched at most once. Predicted entities without a matching reference annotation were counted as false positives (FP), whereas reference entities without a matching prediction were counted as false negatives (FN).

Precision, recall and F1-score were then computed as:

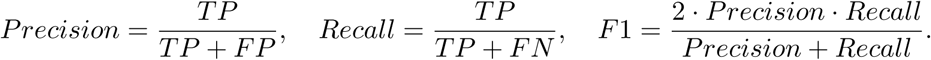

Since true negatives are not defined in span-based NER evaluation, accuracy was calculated as an entity-level agreement score over the union of matched and unmatched annotations:

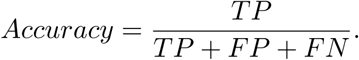

These metrics were used to quantify entity-level agreement between projected and silver-standard annotations in the generated notes. The resulting scores enabled comparison between the RAG-based setting (E2E) and the non-RAG setting (T2E), with particular emphasis on entity preservation, localisation and recoverability.

## 4 Results

### 4.1 Entity Projection

Overall F1-score performance was consistently higher in E2E than in T2E, indicating that the T2E strategy posed a more demanding entity recognition task. As shown in Figure 2, the best E2E result was obtained in the zero-shot setting with GLM5 (F1-score = 0.915), followed by CS4.6 in the same prompting configuration (F1-score = 0.896). In the few-shot E2E experiments, GLM5 again achieved the highest F1-score (0.891), closely followed by CS4.6 (0.889). Qwen3.5:35b also reached competitive performance in E2E, particularly in the zero-shot condition. The zero-shot to few-shot gradient in E2E was therefore limited and mostly negative for the strongest models, with GLM5 decreasing by −0.024 and CS4.6 by −0.007, whereas GPT5.4 nano showed a small positive change (+0.031) from a lower baseline.

**Figure 2:**
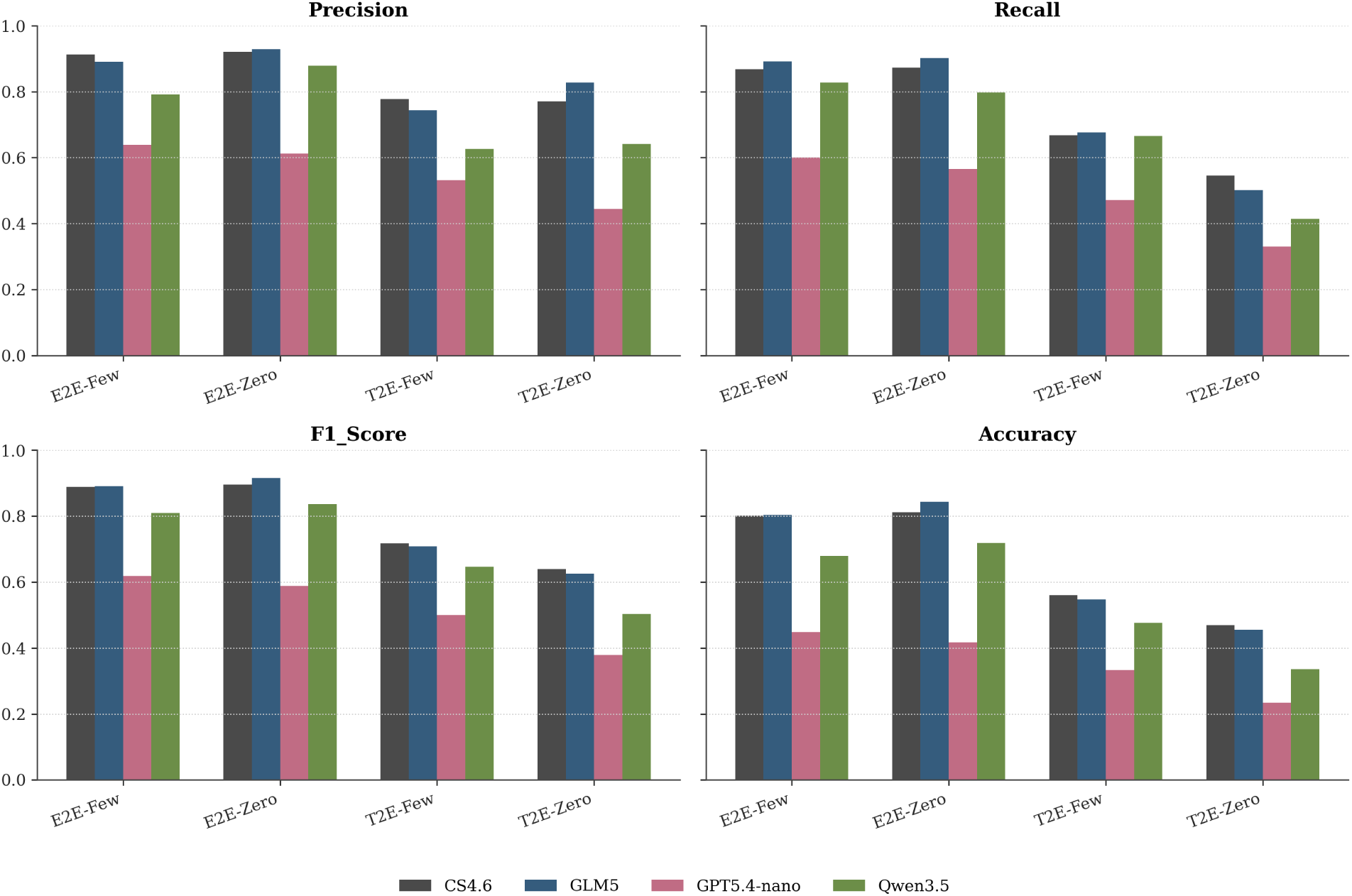
Entity projection metrics across E2E and T2E experiments and models.

In T2E, few-shot prompting produced a clearer improvement over zero-shot prompting. CS4.6 achieved the best performance in both T2E settings, increasing from 0.639 in zero-shot to 0.718 in few-shot (Δ = +0.079). GLM5 followed a similar pattern, reaching 0.708 in T2E few-shot (Δ = +0.083). The largest few-shot gains were observed for Qwen3.5:35b (Δ = +0.143) and GPT5.4 nano (Δ = +0.121), indicating that examples were particularly useful for models that otherwise struggled more with the zero-shot T2E formulation. This suggests that, unlike E2E, T2E benefits substantially from examples that clarify the intended annotation boundaries and label usage.

The entity-level results in Table 3 show that some clinically important labels were recognised robustly across prompting configurations. Age achieved consistently high F1-scores, while *Gleason* was among the best-performing entities in most configurations. This is particularly relevant in the context of prostate cancer, where Gleason score is a key prognostic and stratification variable. *Disease*, *Procedure*, *Duration*, negation-related labels, and *Symptom* also performed strongly in E2E, especially for CS4.6 and GLM5. At entity level, the few-shot versus zero-shot gradients were not uniformly positive in E2E, reinforcing the global pattern that examples did not systematically improve the already well-constrained setting.

**Table 3:** Entity-level F1-score comparison across E2E and T2E generation experiments. Values correspond to F1-scores computed independently for each entity type. The table includes entity types with an aggregate silver-standard support of at least 500 mentions. For each model, the zero-shot setting is used as baseline, and the Δ column reports the difference between few-shot and zero-shot performance. Green indicates improvement and red indicates degradation.

| Entity | CS4.6 |  |  | GLM5 |  |  | GPT5.4 nano |  |  | Qwen3.5 |  |  |
| --- | --- | --- | --- | --- | --- | --- | --- | --- | --- | --- | --- | --- |
| | Zero | Few | $\Delta$ | Zero | Few | $\Delta$ | Zero | Few | $\Delta$ | Zero | Few | $\Delta$ |
| Age | 0.98 | 0.99 | **+0.01** | 0.99 | 0.99 | **0.00** | 0.95 | 0.99 | **+0.04** | 0.96 | 0.98 | **+0.01** |
| Chemicals | 0.93 | 0.92 | **-0.01** | 0.92 | 0.88 | **-0.04** | 0.64 | 0.58 | **-0.06** | 0.86 | 0.83 | **-0.03** |
| Date | 0.86 | 0.86 | **0.00** | 0.84 | 0.86 | **+0.02** | 0.55 | 0.64 | **+0.09** | 0.72 | 0.81 | **+0.09** |
| Disease | 0.91 | 0.90 | **-0.01** | 0.93 | 0.89 | **-0.04** | 0.62 | 0.64 | **+0.02** | 0.88 | 0.82 | **-0.06** |
| Dose | 0.88 | 0.85 | **-0.03** | 0.93 | 0.67 | **-0.26** | 0.78 | 0.45 | **-0.34** | 0.80 | 0.59 | **-0.20** |
| Duration | 0.94 | 0.95 | **0.00** | 0.94 | 0.95 | **0.00** | 0.60 | 0.65 | **+0.05** | 0.88 | 0.88 | **0.00** |
| Frequency | 0.95 | 0.96 | **+0.01** | 0.93 | 0.91 | **-0.02** | 0.77 | 0.62 | **-0.14** | 0.90 | 0.91 | **+0.02** |
| Gleason | 0.98 | 1.00 | **+0.02** | 0.99 | 1.00 | **+0.01** | 0.66 | 0.76 | **+0.10** | 0.86 | 0.99 | **+0.13** |
| Human | 0.87 | 0.94 | **+0.07** | 0.99 | 0.98 | **0.00** | 0.68 | 0.86 | **+0.18** | 0.27 | 0.93 | **+0.66** |
| Neg_cue | 0.93 | 0.94 | **+0.02** | 0.93 | 0.94 | **+0.01** | 0.53 | 0.58 | **+0.05** | 0.86 | 0.89 | **+0.04** |
| Negated | 0.86 | 0.89 | **+0.03** | 0.88 | 0.87 | **-0.01** | 0.52 | 0.59 | **+0.08** | 0.80 | 0.86 | **+0.06** |
| PSA | 0.98 | 0.77 | **-0.20** | 0.99 | 0.88 | **-0.11** | 0.60 | 0.64 | **+0.04** | 0.94 | 0.17 | **-0.77** |
| Procedure | 0.90 | 0.86 | **-0.04** | 0.91 | 0.90 | **-0.01** | 0.57 | 0.61 | **+0.05** | 0.86 | 0.81 | **-0.05** |
| Proteins | 0.93 | 0.95 | **+0.02** | 0.94 | 0.93 | **-0.01** | 0.58 | 0.62 | **+0.04** | 0.88 | 0.87 | **-0.01** |
| Spec_cue | 0.84 | 0.92 | **+0.08** | 0.94 | 0.92 | **-0.03** | 0.52 | 0.55 | **+0.04** | 0.84 | 0.87 | **+0.02** |
| Species | 0.91 | 0.87 | **-0.04** | 0.92 | 0.91 | **0.00** | 0.65 | 0.60 | **-0.05** | 0.71 | 0.62 | **-0.10** |
| Speculated | 0.83 | 0.91 | **+0.08** | 0.85 | 0.87 | **+0.01** | 0.52 | 0.59 | **+0.07** | 0.74 | 0.86 | **+0.12** |
| Symptom | 0.85 | 0.83 | **-0.01** | 0.86 | 0.82 | **-0.04** | 0.57 | 0.58 | **0.00** | 0.79 | 0.72 | **-0.07** |

(b) Text-to-Entity (T2E) Generation
| Entity | CS4.6 |  |  | GLM5 |  |  | GPT5.4 nano |  |  | Qwen3.5 |  |  |
| --- | --- | --- | --- | --- | --- | --- | --- | --- | --- | --- | --- | --- |
| | Zero | Few | $\Delta$ | Zero | Few | $\Delta$ | Zero | Few | $\Delta$ | Zero | Few | $\Delta$ |
| Age | 1.00 | 0.99 | **-0.01** | 1.00 | 0.99 | **-0.01** | 0.74 | 1.00 | **+0.26** | 0.97 | 0.96 | **-0.01** |
| Chemicals | 0.73 | 0.74 | **+0.02** | 0.73 | 0.66 | **-0.07** | 0.41 | 0.34 | **-0.07** | 0.55 | 0.51 | **-0.05** |
| Date | 0.72 | 0.74 | **+0.01** | 0.68 | 0.72 | **+0.04** | 0.45 | 0.52 | **+0.07** | 0.52 | 0.69 | **+0.16** |
| Disease | 0.59 | 0.69 | **+0.10** | 0.66 | 0.71 | **+0.06** | 0.39 | 0.45 | **+0.06** | 0.48 | 0.63 | **+0.16** |
| Dose | 0.46 | 0.31 | **-0.15** | 0.64 | 0.30 | **-0.35** | 0.38 | 0.39 | **+0.01** | 0.36 | 0.24 | **-0.12** |
| Duration | 0.53 | 0.80 | **+0.27** | 0.27 | 0.79 | **+0.52** | 0.21 | 0.58 | **+0.37** | 0.78 | 0.76 | **-0.02** |
| Frequency | 0.81 | 0.85 | **+0.04** | 0.73 | 0.68 | **-0.05** | 0.36 | 0.54 | **+0.18** | 0.46 | 0.65 | **+0.19** |
| Gleason | 0.95 | 1.00 | **+0.05** | 0.99 | 0.98 | **-0.01** | 0.18 | 0.80 | **+0.61** | 0.23 | 0.96 | **+0.73** |
| Human | 0.62 | 0.92 | **+0.30** | 0.09 | 0.92 | **+0.83** | 0.00 | 0.90 | **+0.90** | 0.00 | 0.95 | **+0.95** |
| Neg_cue | 0.79 | 0.85 | **+0.07** | 0.65 | 0.82 | **+0.17** | 0.42 | 0.43 | **+0.01** | 0.06 | 0.74 | **+0.68** |
| Negated | 0.72 | 0.78 | **+0.07** | 0.57 | 0.76 | **+0.19** | 0.28 | 0.45 | **+0.18** | 0.00 | 0.67 | **+0.67** |
| PSA | 0.82 | 0.41 | **-0.41** | 0.84 | 0.24 | **-0.60** | 0.63 | 0.54 | **-0.09** | 0.84 | 0.25 | **-0.59** |
| Procedure | 0.75 | 0.77 | **+0.02** | 0.72 | 0.79 | **+0.07** | 0.47 | 0.57 | **+0.10** | 0.61 | 0.76 | **+0.15** |
| Proteins | 0.46 | 0.81 | **+0.36** | 0.30 | 0.78 | **+0.48** | 0.27 | 0.65 | **+0.38** | 0.48 | 0.68 | **+0.20** |
| Spec_cue | 0.54 | 0.82 | **+0.28** | 0.54 | 0.76 | **+0.22** | 0.13 | 0.22 | **+0.09** | 0.17 | 0.67 | **+0.49** |
| Species | 0.30 | 0.29 | **-0.01** | 0.41 | 0.48 | **+0.07** | 0.04 | 0.14 | **+0.10** | 0.15 | 0.00 | **-0.15** |
| Speculated | 0.58 | 0.81 | **+0.23** | 0.60 | 0.79 | **+0.19** | 0.12 | 0.23 | **+0.11** | 0.00 | 0.54 | **+0.54** |
| Symptom | 0.48 | 0.56 | **+0.08** | 0.50 | 0.53 | **+0.03** | 0.38 | 0.41 | **+0.02** | 0.50 | 0.53 | **+0.03** |

Entity-level performance was more variable in T2E. Few-shot prompting improved several labels that require more contextual interpretation, including *Duration*, *Human*, *Proteins*, *Spec_cue*, and *Speculated*. These gains indicate that examples are useful when the model must infer not only the entity span, but also the annotation convention behind the label. Conversely, *PSA* and *Dose* labels showed unstable behaviour, with marked drops in some few-shot comparisons. This suggests that numerical entities are especially sensitive to the quality and consistency of the examples provided, and that a positive global few-shot gradient can still coexist with label-specific degradation.

### 4.2 LLM-as-a-Judge Evaluation

The LLM-as-a-judge evaluation performed with Kimi K2.6 showed high perceived clinical quality across most model–setting combinations. As shown in Figure 3, mean scores were consistently over 4.5, indicating that most generated notes were judged as clinically coherent, well organised and useful. The strongest overall ratings were obtained by CS4.6, particularly in E2E, although high scores were also observed in several T2E configurations. This indicates that the more demanding entity-level behaviour observed in T2E did not necessarily translate into poor document-level clinical quality.

**Figure 3:**
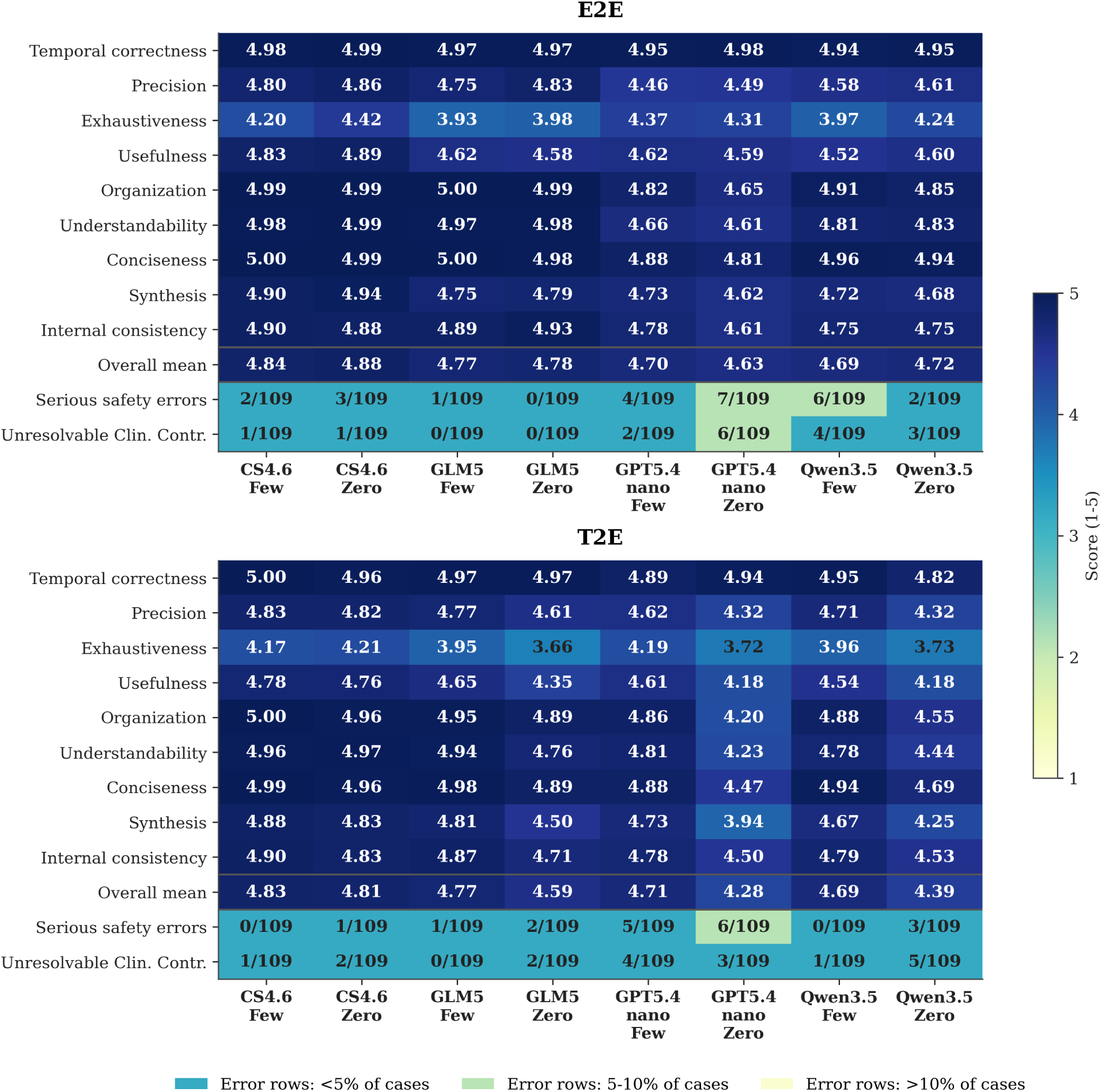
Kimi K2.6 LLM-as-a-judge evaluation of E2E and T2E synthetic clinical progress notes. Heatmap values show the mean score across 109 cases for each model and prompting strategy. The two bottom rows report serious safety errors and unresolvable clinical contradictions as counts over 109 cases, with colour intensity reflecting predefined error-rate bands: <5%, 5-10%, and >10% of cases.

The comparison between E2E and T2E suggests that perceived note quality and entity-level fidelity capture different aspects of performance. While T2E produced lower entity-level F1-scores, several T2E outputs retained high judge-based ratings. This indicates that generated notes may remain readable, clinically plausible and internally coherent even when their annotations are less stable at span level. Therefore, the LLM-as-a-judge evaluation provides complementary information to the entity projection metrics, particularly for assessing global clinical adequacy, document organisation and safety-related issues.

Safety-critical errors and irreconcilable contradictions were infrequent overall, but they were concentrated in specific model–setting combinations. The heatmap of Figure 3 shows that GPT5.4 nano accumulated the highest number of safety-related flags in several settings, whereas GLM5 and CS4.6 generally showed more stable safety profiles. Qwen3.5:35b also presented isolated configurations with relevant contradiction counts. These results indicate that high average judge-based scores can coexist with clinically relevant failure modes, reinforcing the need to analyse safety events separately from global quality ratings.

Few-shot prompting had a heterogeneous effect. In T2E, it generally improved judge-based quality for several models, consistent with the entity-level results showing that this setting benefited from additional examples. However, few-shot prompting did not fully eliminate safety-relevant failures. This indicates that examples may improve global structure and task alignment while still failing to prevent errors involving polarity, numerical interpretation or clinically critical statements.

### 4.3 Error Analysis

A case-level inspection of flagged outputs showed that the most relevant failures were not limited to omissions, formatting problems or stylistic deviations. Instead, several errors involved clinically meaningful changes in polarity, negation, numerical interpretation or contextual meaning. These failures are particularly relevant because they may preserve the overall plausibility of the generated note while altering clinically salient information from the source case.

Case 113 illustrates this type of failure, as shown in Figure D1 in the Appendix D. In the source case, the tumour was reported as positive for PSAP, PSA and PAS, and negative for CEA. Several generated notes altered this interpretation, producing immunohistochemical descriptions that were inconsistent with the original case. In prostate cancer documentation, this type of polarity error is relevant because marker status contributes to the interpretation of tumour origin, diagnostic consistency and follow-up logic.

Case 46 concentrated several safety and contradiction flags across different configurations. The affected outputs showed errors involving surgical margin interpretation, deep venous thrombosis status, haemodynamic condition and postoperative complications. Although these errors differed in their specific formulation, they shared a common pattern: clinically relevant information was inverted, made ambiguous or rendered internally inconsistent. Such errors are particularly important because they may affect oncological follow-up, postoperative risk assessment or therapeutic interpretation.

Overall, the error analysis suggests that the flagged outputs reflect both the sensitivity of certain clinical variables and the heterogeneity of the source material. Differences between Peninsular and Latin American Spanish, older case reports and variable reporting styles may complicate interpretation, particularly for pathology markers, PSA-related expressions, surgical descriptions and negation-dependent statements. At the same time, these categories require precise preservation during generation and projection, making them natural targets for additional normalisation, rule-based validation or expert-in-the-loop review.

These findings also indicate that aggregate entity-level metrics and judge-based quality scores should be complemented with case-level error analysis. A generated note may obtain high global quality scores and still contain local errors affecting clinically decisive statements. Therefore, future entity-aware generation systems should incorporate targeted validation mechanisms for polarity, negation, numerical values, biomarker status and other high-impact clinical variables.

## 5 Discussion

The results suggest that entity-level performance was influenced by both model capacity and the degree of constraint imposed by each generation setting. E2E provided a more clearly delimited scenario, in which the strongest models achieved high F1-scores without requiring substantial benefit from few-shot examples. GLM5 and CS4.6 obtained the best overall results, while Qwen3.5:35b remained competitive, suggesting that locally deployable models can perform adequately when the task is sufficiently constrained and the expected annotation behaviour is explicit.

T2E showed a different pattern. In this configuration, few-shot prompting produced consistent improvements across models, indicating that examples were more useful when entity boundaries and label interpretation were less directly recoverable from the generated text. This effect was particularly visible for contextual labels such as *Duration*, *Spec_cue*, *Speculated* and *Human*, whose correct annotation depends not only on lexical form but also on the surrounding context.

The results for prostate-cancer-specific entities were also informative. *Gleason* achieved consistently strong performance, which is clinically relevant given its central role in prostate cancer diagnosis, prognosis and risk stratification. This behaviour is likely explained by the relatively regular structure of *Gleason* mentions, which commonly combine a restricted lexical anchor with recognisable numerical patterns. By contrast, *PSA* and *Dose* labelling was more unstable, mainly because of over-annotation. These entities require numerical values to be interpreted together with units, measurement conventions and local clinical context. When units or explicit anchors are omitted—as often occurs in clinically compressed writing—, models may generalise the annotation pattern too broadly and mark isolated numbers as entities. This indicates that quantitative clinical entities are particularly vulnerable to context loss during model-based annotation.

The LLM-as-a-judge results complement the entity-level evaluation by showing that document-level clinical plausibility and span-level annotation fidelity are related but not equivalent. Several configurations achieved high judge-based quality scores despite lower entity-level F1-scores, especially in T2E. This suggests that a synthetic progress note may be coherent, readable and clinically plausible while still being less reliable as a structured resource for entity projection. Consequently, both perspectives are necessary: entity-level metrics quantify preservation and localisation of clinical mentions, whereas judge-based evaluation captures broader aspects of clinical coherence, usefulness and safety.

The safety analysis further shows that clinical reliability cannot be inferred from average quality scores alone. Although serious errors and irreconcilable contradictions were uncommon, they clustered in specific model–setting combinations and affected clinically relevant information. This is particularly important in prostate cancer documentation, where small changes in pathology markers, PSA-related statements, surgical margins or postoperative complications can substantially alter the clinical interpretation of the case.

These findings also qualify the role of few-shot prompting. Additional examples improved performance in several T2E configurations, but they did not fully prevent safety-relevant failures. This suggests that few-shot prompting can improve task alignment and output structure, but it is not sufficient by itself to guarantee clinical correctness. In particular, errors involving contrariness, implicit numerical context and clinically compressed expressions require targeted validation mechanisms beyond average-score evaluation.

Taken together, the results support the feasibility of generating clinically plausible synthetic prostate cancer evolution notes with projected entities, but also indicate that reliability must be assessed at multiple levels. High judge-based scores reflect strong document-level quality, whereas entity-level metrics and error analysis reveal whether the generated text is suitable for downstream information extraction.

### 5.1 Limitations

This study should be interpreted in light of the methodological and computational cost of the experimental design. Although the corpus comprised 109 prostate cancer cases, the complete evaluation involved four generation settings and four models, resulting in a substantially larger number of generated clinical notes and entity-projection outputs to analyse. This design enabled a controlled comparison across models and prompting strategies, but also constrained the feasibility of expanding the dataset further within the available computational, annotation and evaluation resources.

Another consideration is that the entity layer used for projection and comparison was automatically generated and should therefore be understood as a silver-standard annotation rather than a fully expert-curated reference, which would require a substantial investment of highly-trained specialist hours. In addition, the source documents were published clinical case reports rather than real EHR notes. While this choice allowed the use of publicly accessible clinical narratives and avoided the privacy restrictions associated with real patient records, it may not fully capture the variability, incompleteness, abbreviations and institution-specific style of routine hospital documentation.

Finally, some configurations still produced safety-relevant inconsistencies, especially in relation to numerical values, contextual interpretation, polarity and entity omission. These issues were not dominant across all models or prompting configurations, but they indicate that generation quality cannot be assessed only through global similarity or clinical readability. For this reason, synthetic clinical note generation remains dependent on multi-level evaluation strategies that combine clinical judgement, entity-level analysis and targeted error detection such as LLM-as-a-judge or expert evaluation.

## 6 Conclusion

This study evaluated the automatic projection of clinically relevant entities onto LLM-generated synthetic Spanish prostate cancer evolution notes. By comparing two generation settings—E2E, with XML-annotated source cases used as RAG context; and T2E, without pre-annotated input—, we analysed not only the apparent clinical quality of the generated notes, but also their suitability as structured resources for downstream clinical NLP tasks. The results show that synthetic clinical progress notes can preserve a substantial proportion of clinically meaningful information from prostate cancer case reports while supporting entity-level annotation transfer.

Overall, the experiments indicate that larger-capacity models achieved better and more stable results. GLM5 and Claude Sonnet 4.6 obtained the strongest entity-level performance and generally showed more robust document-level behaviour, particularly in the more demanding settings. Although Qwen 3.5 remained competitive in constrained configurations, especially in E2E, the results suggest that model capacity becomes more important when the task requires simultaneous clinical reformulation, entity selection, annotation consistency and preservation of numerical or context-dependent information. This was especially evident in T2E, where the absence of annotated input increased variability and made few-shot examples more useful.

E2E obtained particularly strong results and represents the most reliable configuration in this study. However, its performance should not be interpreted as a simple consequence of providing labelled entities in the original case. Even when the source text includes XML-style annotations as RAG support, the model must still transform a published case report into a concise hospital-style evolution note. This requires deciding which entities should be preserved, reformulated, compressed or discarded. In this sense, entity removal is not necessarily an error: some entities present in the original case report may be irrelevant, redundant or unsuitable for the target clinical note style. The good performance of E2E therefore suggests that annotated contextual support can help models preserve key medical information while still filtering entities that are not useful for the transformation of the text.

The joint analysis of entity-level metrics and LLM-as-a-judge evaluation highlights that clinical adequacy and annotation fidelity capture complementary aspects of generation quality. A note may be clinically coherent, useful and stylistically appropriate while still showing imperfect agreement at the span or entity level. This was particularly visible in some T2E outputs, where the generated texts were judged as clinically acceptable despite lower entity-level alignment. These results suggest that synthetic clinical note generation should not be evaluated through a single criterion, but rather through a combined framework including clinical quality assessment, entity projection performance and targeted analysis of critical inconsistencies.

Future work should extend the evaluation to larger and more heterogeneous clinical datasets, including additional oncology domains and, where feasible, real-world EHR documentation under appropriate governance and privacy frameworks. Further research should also incorporate broader expert validation and develop post-generation verification modules focused on numerical consistency, negation, temporality, polarity and clinically relevant entity preservation. These extensions would help determine whether annotation-aware strategies such as E2E can generalise beyond the current corpus and support the creation of more reliable synthetic resources for clinical NLP.

## Data and Code Availability

Access credentials can be provided upon request for editors wishing to inspect the detailed results (including all generated notes, full entity-level metrics, LLM-as-a-judge evaluations, etc.) in the deployed app CoLLMo Web App. All 109 clinical cases can be found in Dataset, while generated notes can be found in Generated Notes. For replicability purposes, all developed code has been uploaded to the following GitHub repository: Synthetic GenEHR and Projection. Please be aware that code execution requires high computational resources or a paid subscription.

## Acknowledgments

The authors acknowledge the support from the Ministerio de Ciencia e Innovación (MICINN) under project PID2024-155334OB-I00.

### A Appendix Entity definitions and examples

This appendix provides brief definitions of the entity types used in the entity-level evaluation. Examples are translated into English from detected spans in the annotated clinical texts. For traceability, each example reports the NER model output, case number, and experiment from which the detected span was extracted.

**Age** Mentions of patient age or age-related expressions.

- *Example:* “66 years old”.
- *Source:* MedSpaNER xlm-roberta-large-spanish-trials-cases-temp-ent, case 1, E2E few-shot.

**Chemicals** Mentions of pharmacological substances, compounds, biochemical substances, or related biomedical chemicals that can be normalised.

- *Example:* “acetylsalicylic acid”.
- *Source:* PharmaCoNER bsc-bio-ehr-es-pharmaconer, case 30, E2E few-shot.

**Date** Calendar dates or relative temporal references that locate a clinical event in time.

- *Example:* “16th postoperative day”.
- *Source:* MedSpaNER xlm-roberta-large-spanish-trials-cases-temp-ent, case 63, E2E few-shot.

**Disease** Mentions of diseases, diagnoses, pathological conditions, or clinically established disorders.

- *Example:* “vertebral metastases”.
- *Source:* DisTEMIST bsc-bio-ehr-es-distemist, case 98, E2E few-shot.

**Dose** Quantified medication doses or administered amounts, usually including a numerical value and unit.

- *Example:* “11.25 mg, 2 doses”.
- *Source:* MedSpaNER roberta-es-clinical-trials-cases-medic-attr, case 56, E2E few-shot.

**Duration** Expressions indicating the length of time over which a condition, treatment, symptom, or clinical process occurred.

- *Example:* “approximately one year”.
- *Source:* MedSpaNER xlm-roberta-large-spanish-trials-cases-temp-ent, case 83, E2E few-shot.

**Frequency** Expressions describing how often an event, symptom, treatment, or clinical action occurs.

- *Example:* “twice per week”.
- *Source:* MedSpaNER xlm-roberta-large-spanish-trials-cases-temp-ent, case 1, E2E few-shot.

**Gleason** Mentions of Gleason score or Gleason grading patterns used in prostate cancer assessment.

- *Example:* “Gleason 10 (5+5)”.
- *Source:* rule-based Gleason extractor, case 112, E2E few-shot.

**Human** Mentions identifying the patient or another human participant in the clinical narrative.

- *Example:* “male patient”.
- *Source:* LivingNER bsc-bio-ehr-es-livingner-humano, case 24, E2E few-shot.

**Neg_cue** Linguistic cues that introduce or signal negation in the clinical text.

- *Example:* in the phrase “without neoplastic involvement”, the cue is “without”.
- *Source:* MedSpaNER roberta-es-clinical-trials-cases-neg-spec, case 1, E2E few-shot.

**Negated** Clinical entities or findings explicitly stated as absent, excluded, or ruled out within their textual context.

- *Example:* in the phrase “without neoplastic involvement”, the negated entity is “neoplastic involvement”.
- *Source:* MedSpaNER roberta-es-clinical-trials-cases-neg-spec, case 1, E2E few-shot.

**PSA** Mentions of prostate-specific antigen values, measurements, or references to PSA as a clinical marker.

- *Example:* “244.10 ng/ml”.
- *Source:* rule-based PSA extractor, case 24, E2E few-shot.

**Procedure** Mentions of diagnostic, therapeutic, surgical, or radiological procedures.

- *Example:* “adjuvant radiotherapy”.
- *Source:* MedProcNER bsc-bio-ehr-es-medprocner, case 65, E2E few-shot.

**Proteins** Mentions of proteins, biomarkers, or protein-based markers relevant to clinical interpretation.

- *Example:* “androgen receptors”.
- *Source:* PharmaCoNER bsc-bio-ehr-es-pharmaconer, case 82, E2E few-shot.

**Spec_cue** Linguistic cues that introduce uncertainty, suspicion, compatibility, or possibility.

- *Example:* in the phrase “compatible with osteoblastic metastases”, the cue is “compatible with”.
- *Source:* MedSpaNER roberta-es-clinical-trials-cases-neg-spec, case 1, E2E few-shot.

**Speculated** Clinical entities or findings presented as suspected, possible, compatible with, or not fully confirmed.

- *Example:* in the phrase “compatible with osteoblastic metastases”, the speculated entity is “osteoblastic metastases”.
- *Source:* MedSpaNER roberta-es-clinical-trials-cases-neg-spec, case 1, E2E few-shot.

**Species** Mentions of organisms, microorganisms, or biological species relevant to the clinical case.

- *Example:* “Staphylococcus epidermidis”.
- *Source:* LivingNER bsc-bio-ehr-es-livingner-species, case 205, E2E few-shot.

**Symptom** Mentions of symptoms, signs, clinical manifestations, or patient-reported complaints.

- *Example:* “urinary incontinence”.
- *Source:* SympTEMIST bsc-bio-ehr-es-symptemist, case 41, E2E few-shot.

### B Appendix Prompts

#### B.1 E2E

**Listing 1:**
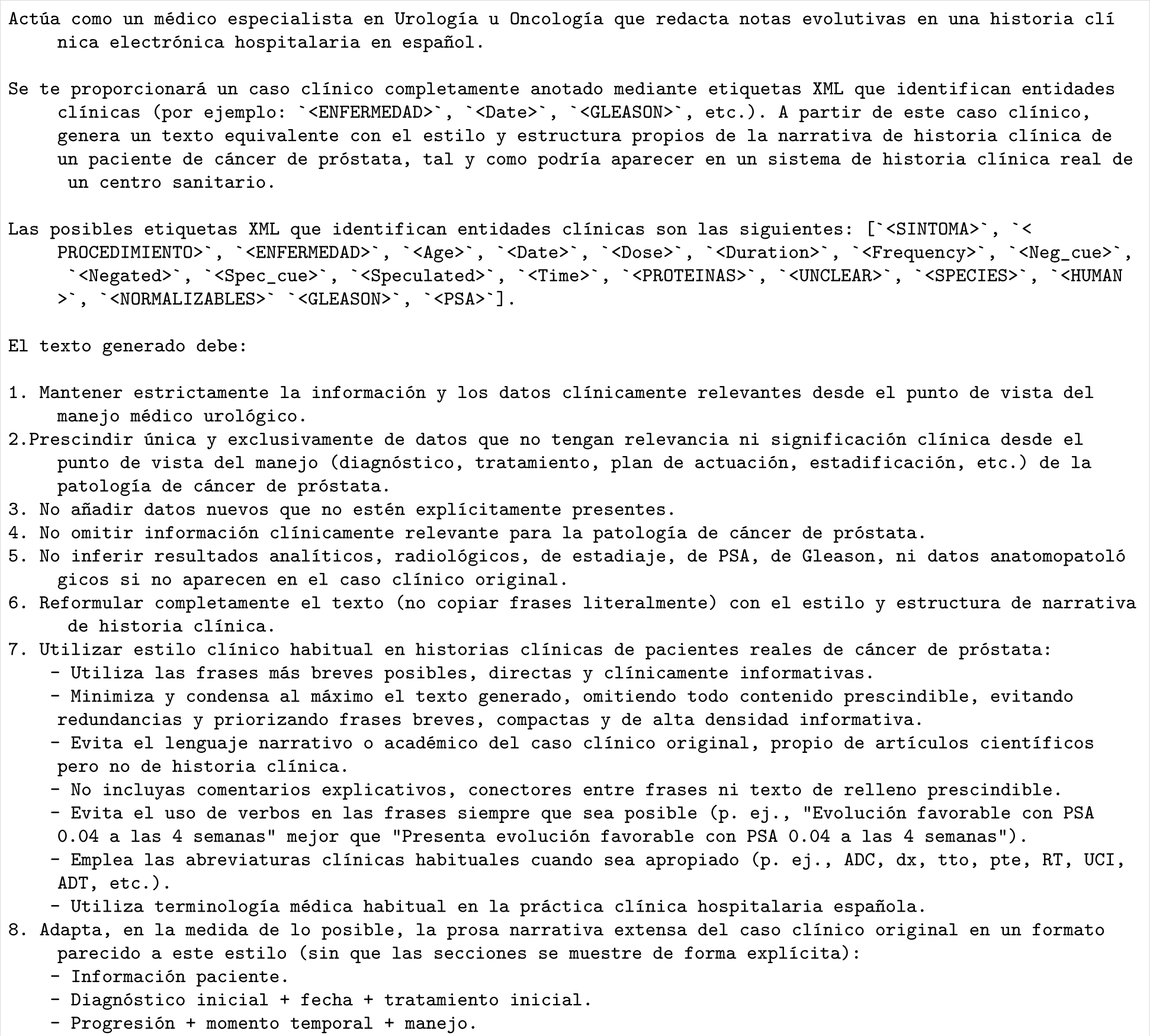
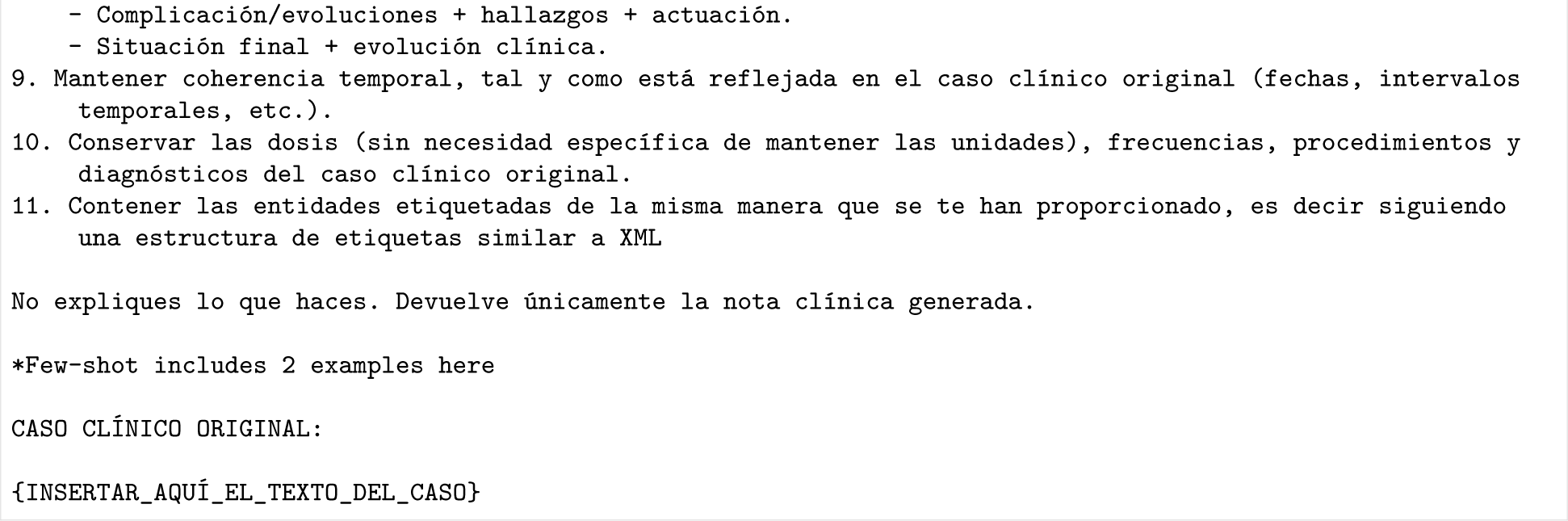
Prompt used for E2E generation

**Listing 2:**
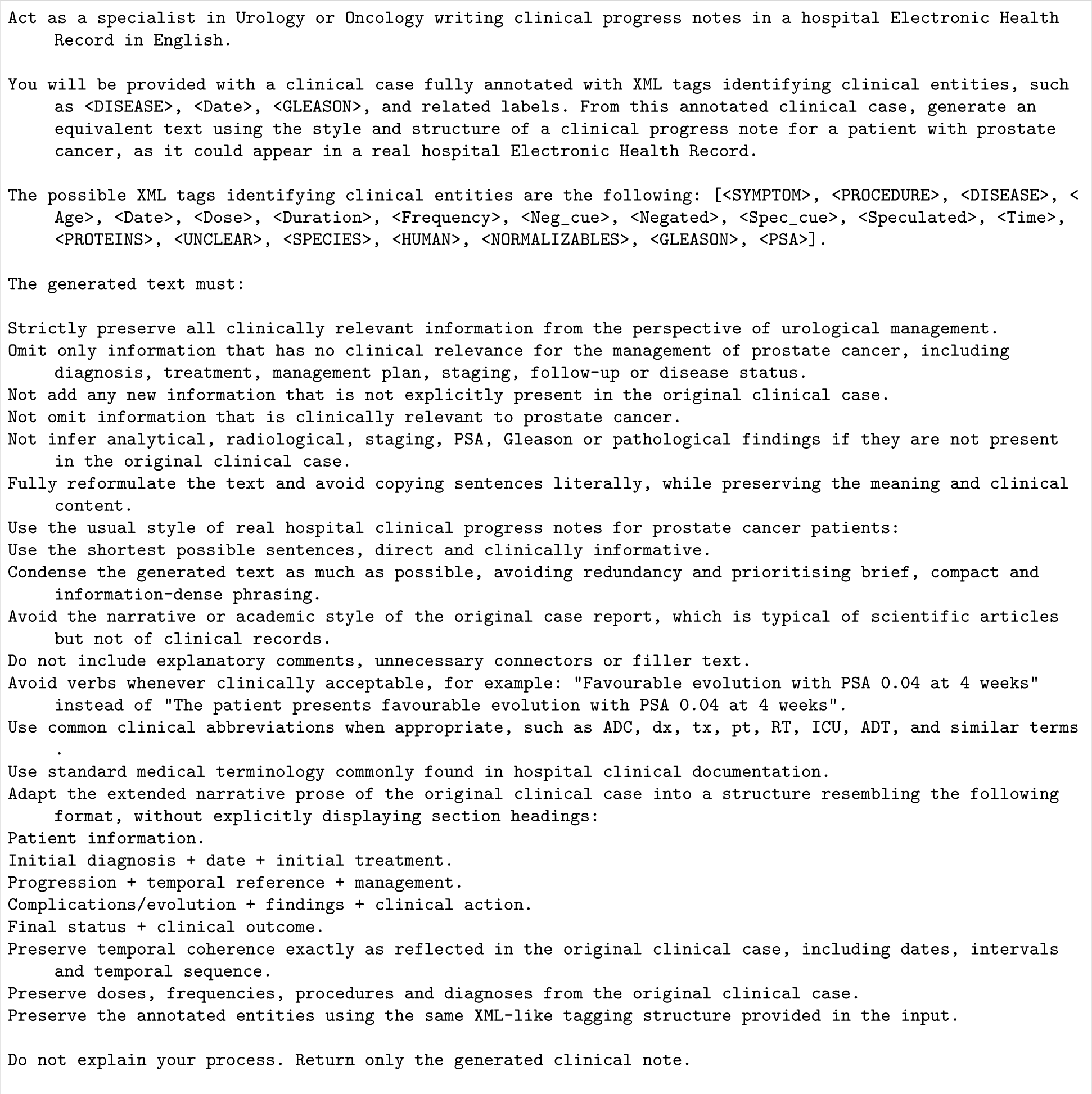
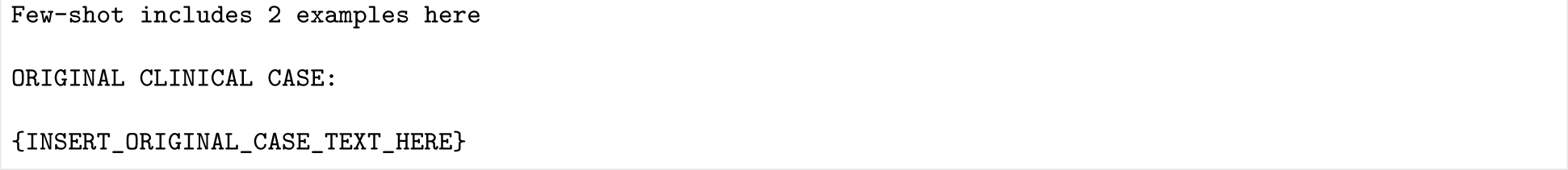
Prompt used for E2E generation in English

#### B.2 T2E

**Listing 3:**
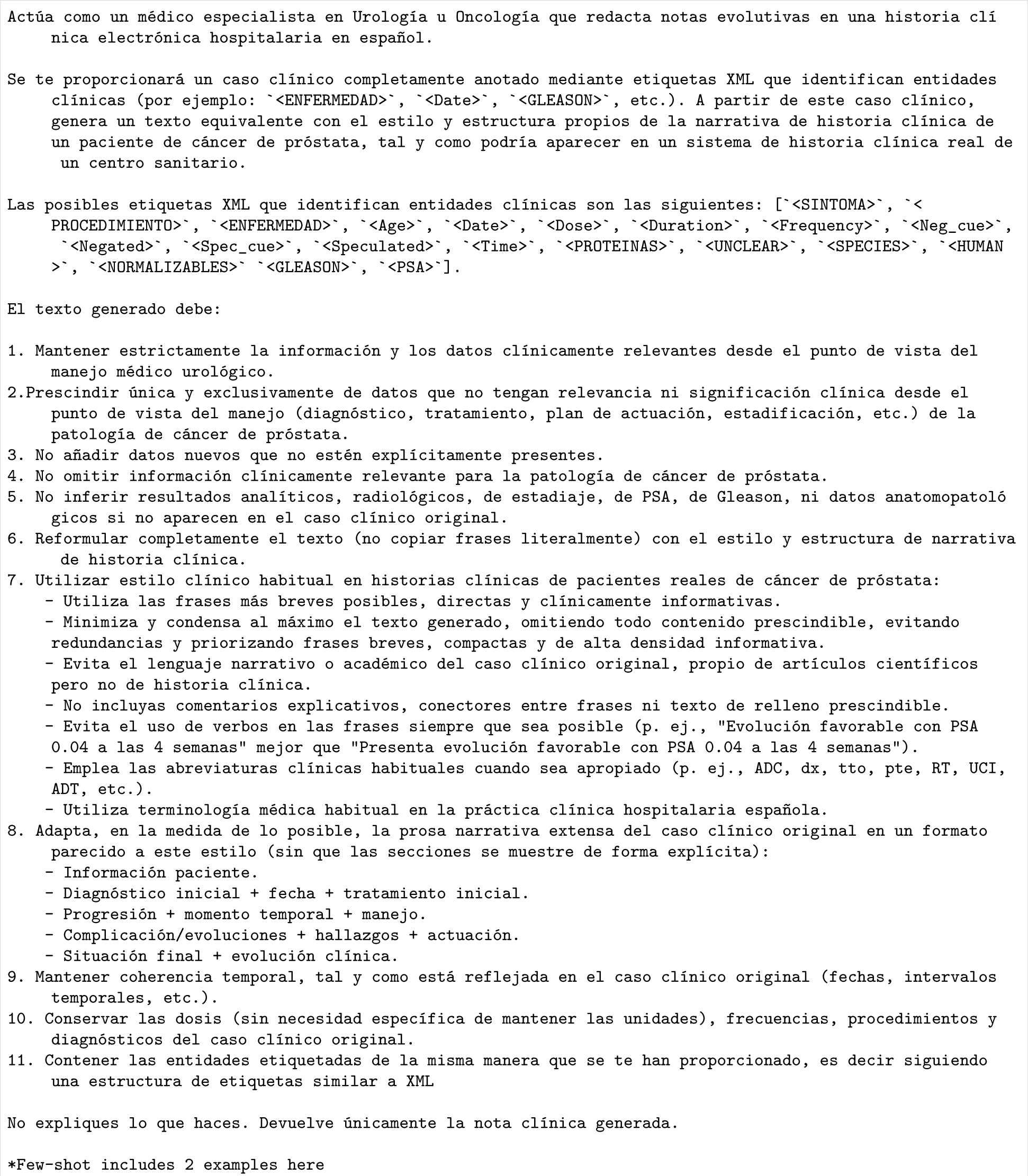
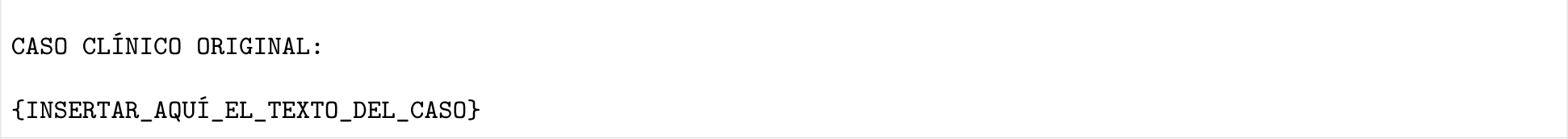
Prompt used for T2E generation

**Listing 4:**
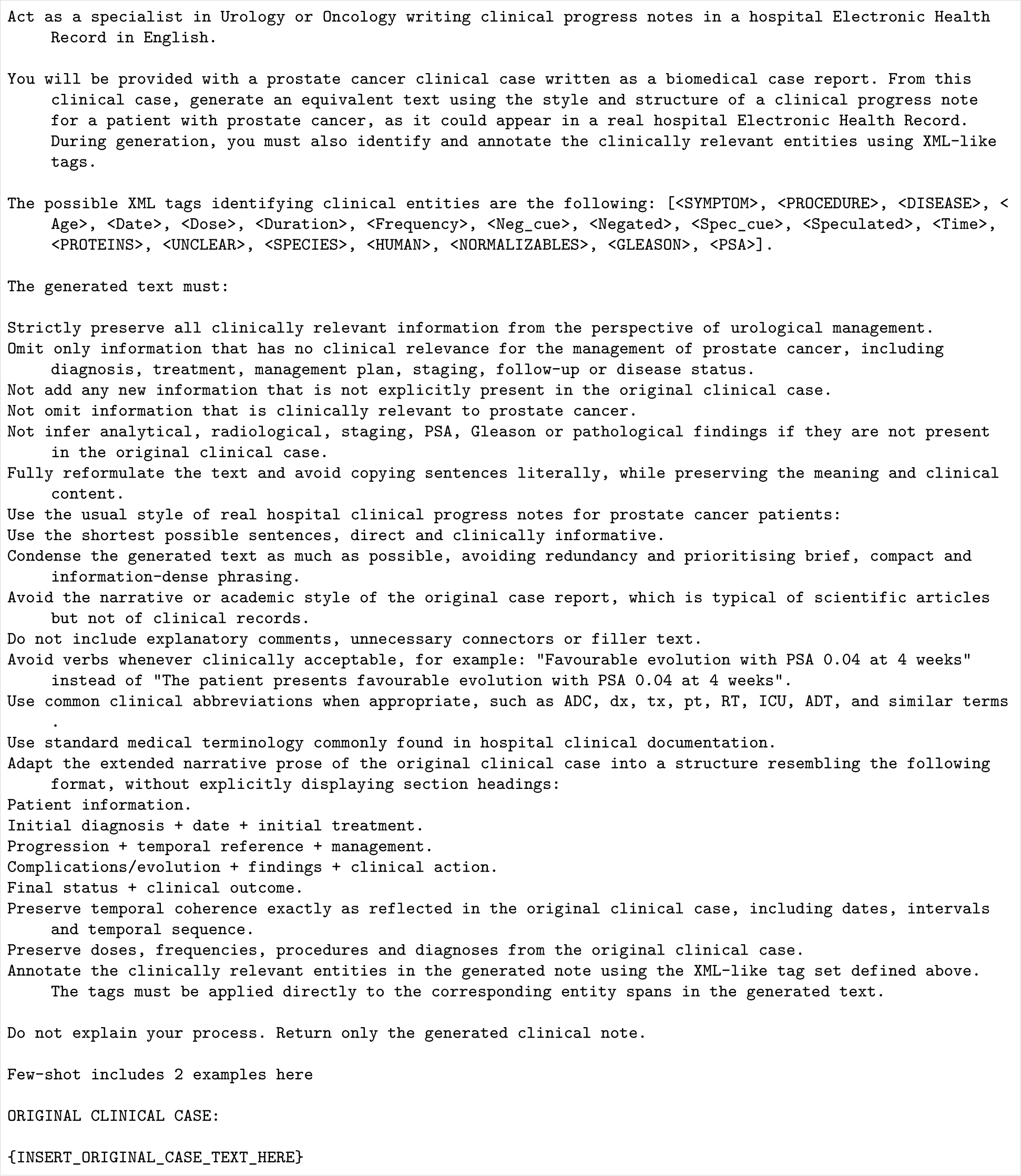
Prompt used for T2E generation in English

### C Appendix Original source case for E2E & T2E

**Figure C1:**
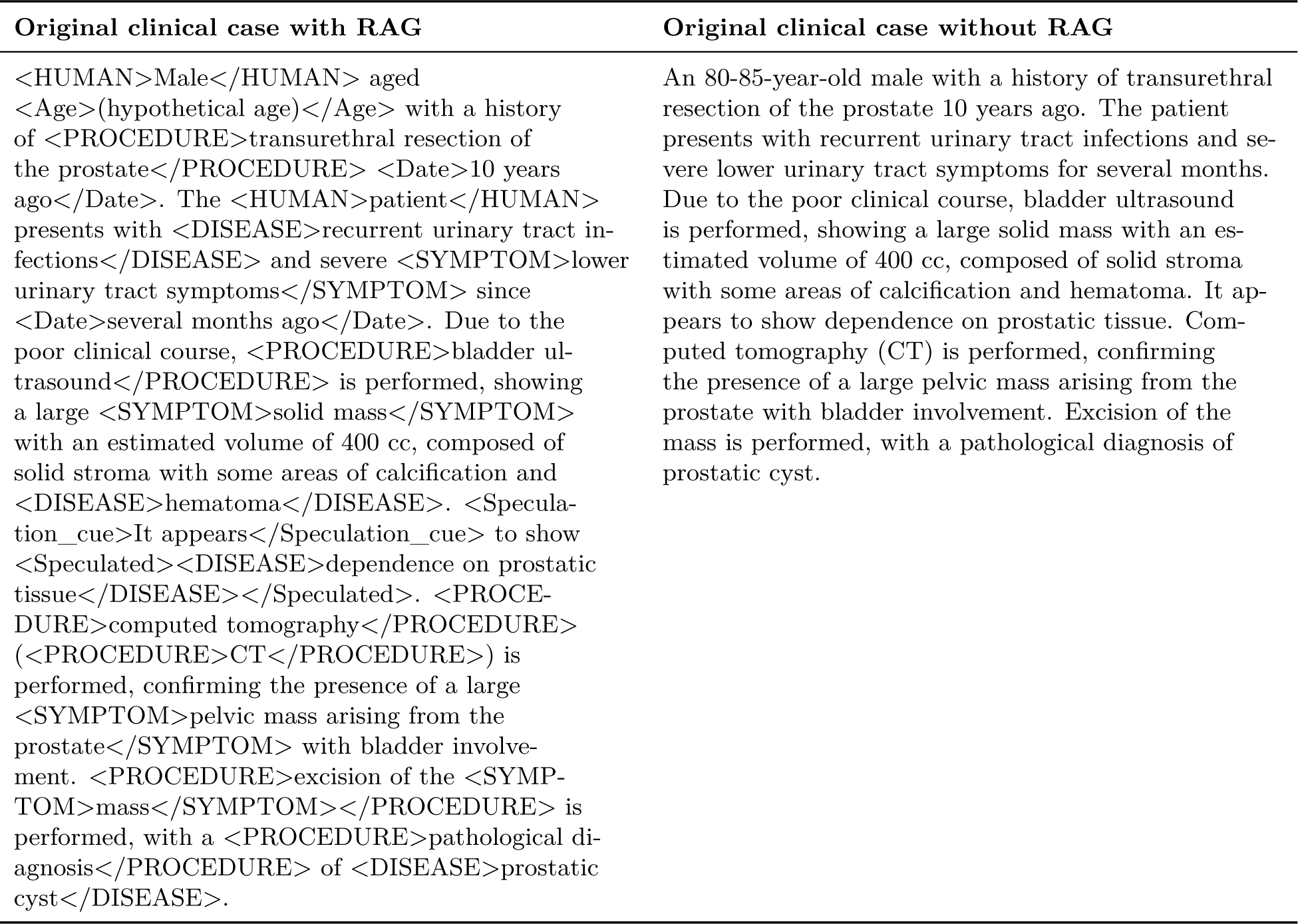
Qualitative example for patient 2 illustrating the main observable difference between E2E and T2E. The left column shows the original clinical case with XML-style entity annotations used by the RAG-based E2E configuration, whereas the right column shows the original clinical case for T2E with the same clinical content expressed as plain text.

### D Error Exemplification

**Figure D1:**
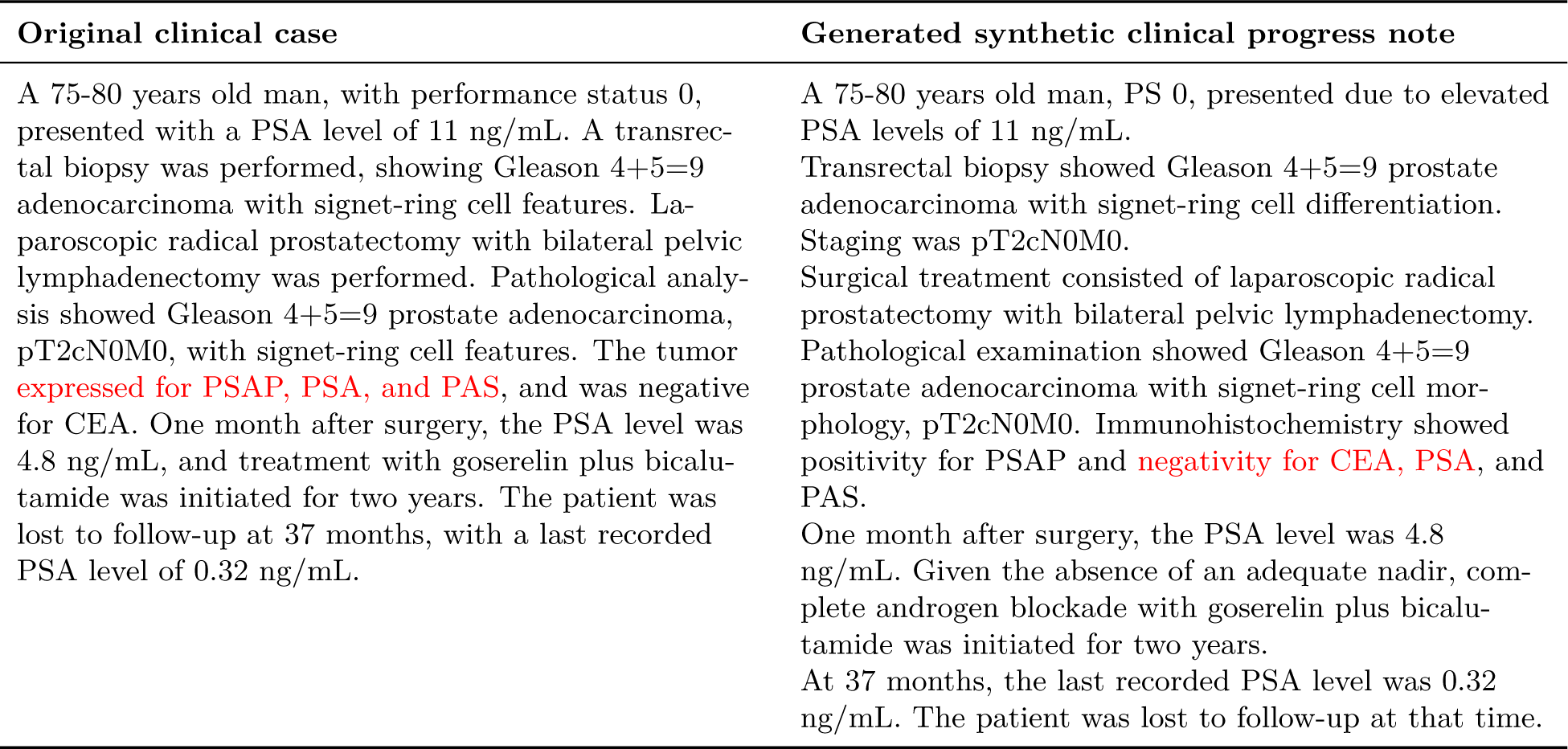
Qualitative example for patient 113 illustrating the main observable safety error. In red, marked error regarding original clinical case report. Generated synthetic clinical progress note corresponds to CS4.6 on E2E zero-shot configuration. Five other experiments—from CS4.6-E2E-few-shot, GLM5-E2E-few, GPT5.4 nano-E2E-zero-shot, Qwen3.5:35b-E2E-few-shot and Qwen3.5:35b-E2E-zero-shot prompting configurations—exhibited the exact same error.

## Footnotes

1 The target narrative style was defined according to the structure, terminology and documentation conventions observed in prostate cancer evolution notes from routine clinical practice at Hospital Universitario Virgen de la Victoria (HUVV), where one of the authors carries out clinical activity. No real patient records were used as input for generation.

